# Epileptic discharges are classified as generalized paroxysmal fast activity with low expert agreement: *findings from an international survey*

**DOI:** 10.64898/2026.09.18.26362075

**Authors:** Sem L. Kampman, Aaron EL. Warren, Itay Tokatly Latzer, Michaela Stamm, John D. Rolston, John S. Archer, Steven Tobochnik, Jurriaan M. Peters

## Abstract

**Objective:** The current International League Against Epilepsy (ILAE) guidelines for Lennox-Gastaut syndrome (LGS) mandate the presence of generalized paroxysmal fast activity (GPFA) on EEG. However, GPFA is not necessarily restricted to LGS and has been described in, among others, idiopathic generalized epilepsies. A clear definition for GPFA is currently lacking, hampering research into its utility as a diagnostic or treatment biomarker. Here, we assessed expert agreement in the interpretation of GPFA to explore feasibility of a consensus-based definition.

**Methods:** We internationally distributed a two-part survey on GPFA to EEG experts. First, participants completed thirteen questions gauging their preferred clinical and EEG criteria for GPFA, including duration, frequency, amplitude and topography. Second, participants classified the presence versus absence of GPFA in eleven EEG clips, while blinded to clinical context, from patients with LGS, other epilepsies, and physiological variants.

**Results:** 162 respondents, predominantly from North America and Europe, completed the survey. 87% considered GPFA to also occur in epilepsies other than LGS. Considerable heterogeneity was present on defining duration, spectral frequency, amplitude and topographic distribution of GPFA. Particularly poor agreement was seen for lower limits of duration and frequency, with no single response option receiving a majority consensus (36% and 30% agreement, respectively). Similarly, the degree of agreement on interpretations was highly variable across real-world EEG clips.

**Significance:** Marked discrepancies exist in the EEG identification and clinical interpretation of GPFA, undermining its utility as a potential biomarker in research and clinical trials of LGS and other epilepsies. The findings also highlight the need to establish a consensus-based and eventually evidence-informed definition to formally operationalize the clinical and EEG criteria for GPFA.

*Key points:*

- Generalized paroxysmal fast activity (GPFA) is variably defined in the literature, limiting its utility as a diagnostic or treatment biomarker
- In this international survey (n = 162 respondents, 25 countries) we found limited agreement on GPFA’s defining features and on interpretation of fast activity on EEG clips
- 87% of respondents considered GPFA to occur in epilepsies other than Lennox-Gastaut syndrome
- Pending further elucidation of the pathophysiology underlying GPFA or its computational characterization (i.e., to inform a biologically-or evidence-based definition), we underscore the need for an interim consensus-based definition

## Introduction

Lennox-Gastaut syndrome (LGS) is a childhood-onset developmental and epileptic encephalopathy defined by multiple drug-resistant seizure types (including tonic seizures and at least one additional seizure type), specific interictal electroencephalography (EEG) features (slow spike-wave [SSW)] and generalized paroxysmal fast activity [GPFA]), and intellectual disability ^1,2^. Diagnosing LGS can be difficult as the exact features necessary for diagnosis have evolved over time and are often applied inconsistently ^3–5^. Diagnosis is further complicated by the shifting nature of EEG abnormalities and seizure types over a patient’s lifetime ^4–9^ and the observation that not all features may be present at epilepsy onset ^1,10^.

According to current International League Against Epilepsy (ILAE) guidelines, the presence of GPFA on either current or historical EEGs is a mandatory criterion for diagnosing LGS ^2^.

However, a standardized definition of GPFA is absent^11,12^. Variable cut-offs for minimum and maximum duration are reported for GPFA, and different electrophysiological characteristics have been reported ^13–15^. The EEG signature of GPFA has been described using varying terms ^16–19^, such as “grand mal pattern” ^20^, “generalized repetitive fast discharges” ^20,21^, “diffuse fast rhythms” ^22^, and “fast rhythmic discharges” ^23^. In clinical settings, terms such as “rapid generalized spikes”, “fast polyspikes” and “diffuse fast beta-alpha range activity” are also commonly encountered. Also, GPFA is not necessarily specific only to LGS ^11,24,25^, having been described in individuals with focal epilepsy ^15,26^ and idiopathic (genetic) generalized epilepsy (IGE) ^14,23,27–33^.

These inconsistencies in the nomenclature and interpretation of GPFA hinder its clinical utility ^11,34^. Increasingly, studies are looking into the role of GPFA as a biomarker of treatment response such as for monitoring seizure burden after changes to medication regimens, brain stimulator settings, or epilepsy surgery ^11,35,36^. However, inaccurate diagnosis and identification of patients with GPFA and/or LGS may reduce the reliability of research findings and limit generalizability to the target population^11^. As such, standardization of GPFA is critical to ensure that the intended patient population is being studied and treatment responses are measured reliably.

Given the ambiguity regarding i) the features that define GPFA, and ii) the clinical interpretation of generalized fast activity– potentially classifiable as GPFA – on EEG, we aimed to quantify specific areas of agreement, using percentage agreement and inter-rater agreement measures.

We asked experts in the field of child and adult neurology to define GPFA, then review and classify EEG clips of fast activity derived from patients with LGS-phenotypes, IGE, focal epilepsies and physiological variants.

## Materials and Methods

The Institutional Review Board (IRB-P00047933) at Boston Children’s Hospital (BCH) approved the project and waived the requirement for informed consent. EEG clips included in the distributed survey were stripped of any identifying information.

### Survey development

We designed a 13-item multiple-choice questionnaire divided into two sections. The first five questions assessed respondent characteristics (e.g., training, practice setting, country of practice, and LGS caseload); next, eight questions assessed the clinical and EEG criteria used when diagnosing GPFA, including duration, frequency, amplitude, topography, and associated epilepsy types. Based on literature review, no questions concerning upper amplitude limit were included; however, a lower limit of amplitude was included as it may help differentiate GPFA from generalized low-voltage fast activity ^28,37^.

In the second part of the survey, respondents were presented with eleven 10-second EEG clips containing fast activity potentially classifiable as GPFA. These included fast rhythms sourced from patients carrying a clinical diagnosis of LGS, as well as clips showing morphologically similar patterns of fast activity from patients with focal epilepsy and IGE. One EEG clip of sleep spindles in a neurotypical patient with non-refractory focal epilepsy served as a “negative control”. Our samples varied in EEG frequency, amplitude and morphology patterns, and in patient characteristics, to explore agreement between respondents across a broad range of electrographic and clinical presentations. Respondents were asked to classify activity as (i) “GPFA”; (ii) “not GPFA, but pathological”; or (iii) “not GPFA, and not pathological”. If they considered the activity “not GPFA, but pathological”, two follow-up questions probed why GPFA criteria was considered not met and what they instead would have classified the EEG activity as. Apart from age and sex, no patient diagnostic information was available to respondents.

Static EEG clips (screenshots) were exported from Natus, with a duration of 10 seconds and time base of 30mm/sec. Low-frequency filters (1 Hz), high-frequency filters (70 Hz) and notch filters (60 Hz) were applied. To allow for assessment of signal amplitude, each EEG clip was shown with both a sensitivity of 7uV/min and 30 uV/min, and a scale legend was provided.

The full survey with all answer options is available in the *Supplemental Material*.

Survey development occurred in three rounds. In Round 1, SK and JP developed the initial survey questionnaire. In Round 2, investigators (SK, AW, ITL, ST, JP) jointly provided feedback and met in April 2024 to finalize the study objectives. In Round 3, full consensus on the final version of the survey was reached (SK, AW, ITL, ST, JP), after which the questions were imported into the BCH REDCap environment, a web-based platform.

### Patient characteristics

Characteristics of patients included in the EEG clips are summarized in *Table 1*. The selection of patients included six patients with multiple LGS features (i.e., childhood onset epilepsy, drug-resistant tonic seizures, multiple seizure types and intellectual disability). Formal diagnostic ILAE criteria for LGS require the presence of both GPFA and SSW on historical or current EEG^2^. Because our study set out to examine whether respondents considered observed fast activity as GPFA, we refrained from assigning a definitive diagnosis in this study; rather, we classified these patients as having an LGS-phenotype. This approach is similar to recent work that used the terms “clinically accepted” or “clinically defined” LGS for patients with incomplete or ambiguous EEG findings (in contrast to patients with “complete electroclinical LGS”)^38^. That study found largely overlapping clinical profiles (including seizure types and treatment resistance) between the “clinically defined LGS” and “complete electroclinical LGS” groups^38^.

**Table 1.** Clinical and demographic characteristics of patients in EEG clips. Apart from sex and age at EEG, these characteristics were not visible to respondents during the survey. *EEG clips #3 and #6 were sourced from the same patient during the same recording. **Note that age at EEG in years was displayed to respondents during the survey, rather than an age range.

| Visible to respondents during survey |  |  | Not visible to respondents during survey |  |  |  |
| --- | --- | --- | --- | --- | --- | --- |
| EEG clip # | Age at EEG (range) ** | Sex | Epilepsy phenotype | Current or historical tonic seizures | Etiology | Intellectual disability |
| 1 | 6-10 | F | LGS-phenotype | Yes | Structural + genetic (stroke + Parry-Romberg syndrome) | Yes |
| 2 | 6-10 | F | Non-refractory focal epilepsy | No | Structural (right parietal encephalomalacia, due to remote stroke) | No |
| 3* | 11-15 | F | LGS-phenotype | Yes | Structural (right parietal encephalomalacia, due to remote MCA stroke) | Yes |
| 4 | 6-10 | M | LGS-phenotype | Yes | Unknown | Yes |
| 5 | 11-15 | F | LGS-phenotype | Yes | Unknown | Yes |
| 6* | 11-15 | F | LGS-phenotype | Yes | Structural (right parietal encephalomalacia, due to remote MCA stroke) | Yes |
| 7 | 6-10 | F | LGS-phenotype | Yes | Structural (congenital stroke) | Yes |
| 8 | 16-20 | F | IGE (refractory JAE) | No | Genetic (presumed, in context of IGE) | No |
| 9 | 16-20 | F | Refractory focal epilepsy | No | Unknown | Yes |
| 10 | 16-20 | F | LGS-phenotype | Yes | Structural (right frontal encephalomalacia, due to subdural empyema) | Yes |
| 11 | 26-30 | M | Refractory focal epilepsy | No | Structural (post right frontal atypical teratoid rhabdoid tumor resection) | Yes |

**Abbreviations.** JAE: juvenile absence epilepsy; MCA: middle cerebral artery.

The other patients included two with refractory focal seizures and intellectual disability, one with medically controlled IGE and normal neurodevelopment, and one patient with non-refractory focal epilepsy with spindles on the EEG (“negative control”).

### Survey distribution

The survey link was published on Child Neurology Society (CNS) and ACNS (American Clinical Neurophysiology Society) online forums, on the American Epilepsy Society (AES) social media platform (X/Twitter) and in the Pediatric Epilepsy Research Consortium (PERC) monthly newsletter. We distributed the survey via email to faculty, residents, and epilepsy fellows at the Division of Epilepsy and Clinical Neurophysiology at BCH and to personal contacts of authors globally. Correspondence information of neurologists, epileptologists and neurophysiologists was retrieved from websites of epilepsy clinics and teaching hospitals, and from publications in epilepsy journals. Lastly, business cards with a QR code linked to the survey were distributed at the 2024 American Epilepsy Society Annual Meeting in Los Angeles, California (USA).

### Study participants

Epileptologists and clinical neurophysiologists, child and adult neurologists and neurology residents, and epilepsy fellows were eligible for participation in the survey. No personal identifying information was collected from respondents; by completing the survey, consent from respondents for participation was inferred. Participation was voluntary, and compensation was not offered to participants.

### Data analysis

Data collection occurred within the REDCap online environment. The survey was online from October 2024 until June 2025 (~ 9 months), whereafter data was exported to R Studio (version 4.5) ^39^ for analysis. We screened data for incomplete responses and, where feasible, reclassified text-based answers (i.e., when a respondent selected ‘*Other’* and provided a textual response) to pre-defined answer options when aligning closely. Summary tables were created to display response counts and percentages for clinical and EEG criteria of GPFA, and respondent interpretations of EEG clips.

For the primary analysis, we used percentage agreement to evaluate agreement among respondents for the EEG clips. An arbitrary majority-based ranking – strong majority (>80% agreement), small majority (≥50%), and no majority (<50%) – was used to simplify comparisons. In the secondary analysis, to evaluate the robustness of the percentage agreement method for probing the degree of agreement, we used Gwet’s AC2’s method for inter-rater agreement with ordinal weighting ^40–42^. This method corrects for chance agreement and facilitates comparison of inter-rater agreement across individual questions. We used Landis and Koch’s scale to interpret Gwet’s AC2: <0.00 = “poor agreement”, 0.00-0.20 = “slight agreement”, 0.21-0.40 = “fair agreement”, 0.41-0.60 = “moderate agreement”, “0.61-0.80” = “substantial agreement”, and 0.81 – 1.00 = “almost perfect agreement” ^43^. Although typically used for kappa statistics^44^, we applied this scale here only to facilitate relative comparisons.

Post hoc, exploratory subgroup analyses were conducted to explore differences in response distributions to the 10-second EEG clips: clinicians with ≤10 vs. 10 years of clinical experience; child vs. adult neurologists; European vs. North American respondents; and clinicians seeing 0-25 vs. 25-50 LGS patients per year. Besides visual review of response distributions, Fisher-exact tests with Holm’s correction for multiple comparisons were performed with statistical significance evaluated at α = 0.05.

## Results

162 respondents completed the survey. Responses were complete for all questions on respondent characteristics and EEG and clinical criteria for GPFA, while only one or two responses were missing for interpretations of several EEG clips. 49% of respondents were child neurologists, and 39% were adult neurologists (*Table 2*). More than half (58%) of respondents had >10 years of clinical experience. 90% practiced in a university-based hospital. Participants were from 25 countries, with North America the most represented continent (57%), followed by Europe (28%).

**Table 2.** Baseline characteristics of respondents (n=162). * “Other” includes clinical neurophysiologists, qualification for more than one category, or ambiguous classification. In cases where respondents clearly indicated both child and adult neurology/epilepsy training, they were classified as child neurologists. If years of experience was not provided in the text section, responses were not re-assigned and instead left as “Other”.

|  | Count | Percentage |
| --- | --- | --- |
| **What is your clinical experience?** |  |  |
| Adult neurology resident or epilepsy fellow/trainee | 4 | 2% |
| Adult neurologist, staff physician (< 5 years experience) | 10 | 6% |
| Adult neurologist, staff physician (5-10 years experience) | 9 | 6% |
| Adult neurologist, staff physician (>10 years experience) | 46 | 28% |
| Child neurology resident or epilepsy fellow/trainee | 4 | 2% |
| Child neurologist, staff physician (< 5 years experience) | 15 | 9% |
| Child neurologist, staff physician (5-10 years experience) | 17 | 10% |
| Child neurologist, staff physician (>10 years experience) | 49 | 30% |
| Other* | 8* | 5% |
| **Do you consider yourself an EEG expert?** |  |  |
| Yes (e.g., dedicated training, board certification in epilepsy or clinical neurophysiology, regular EEG interpretation in clinical setting) | 158 | 98% |
| No | 4 | 2% |
| **In which healthcare setting do you currently work?** |  |  |
| University-based hospital / academic hospital | 145 | 90% |
| Non-university-based hospital | 17 | 10% |
| Private practice | 0 | 0% |
| **Continent currently practicing in** |  |  |
| North America | 92 | 57% |
| Europe | 45 | 28% |
| Asia | 13 | 8% |
| South America | 2 | 1% |
| Oceania | 8 | 5 % |
| Africa | 1 | 1% |
| Eurasia (Turkey) | 1 | 1% |
| **How many patients with LGS do you see per year?** |  |  |
| <10 | 39 | 24% |
| 10 to 25 | 50 | 31% |
| 25 to 50 | 42 | 26% |
| >50 | 31 | 19% |

**Table 3.**
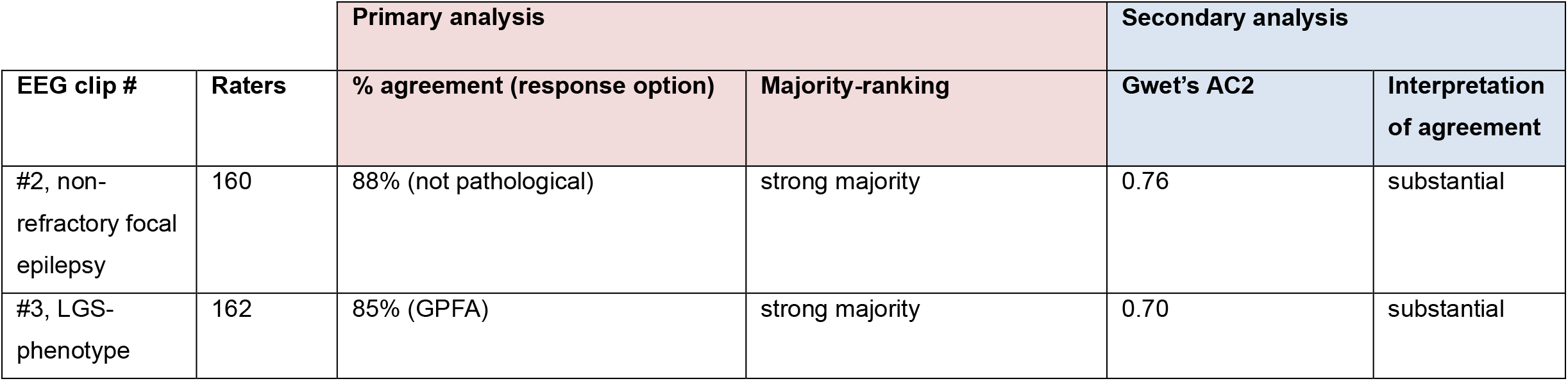
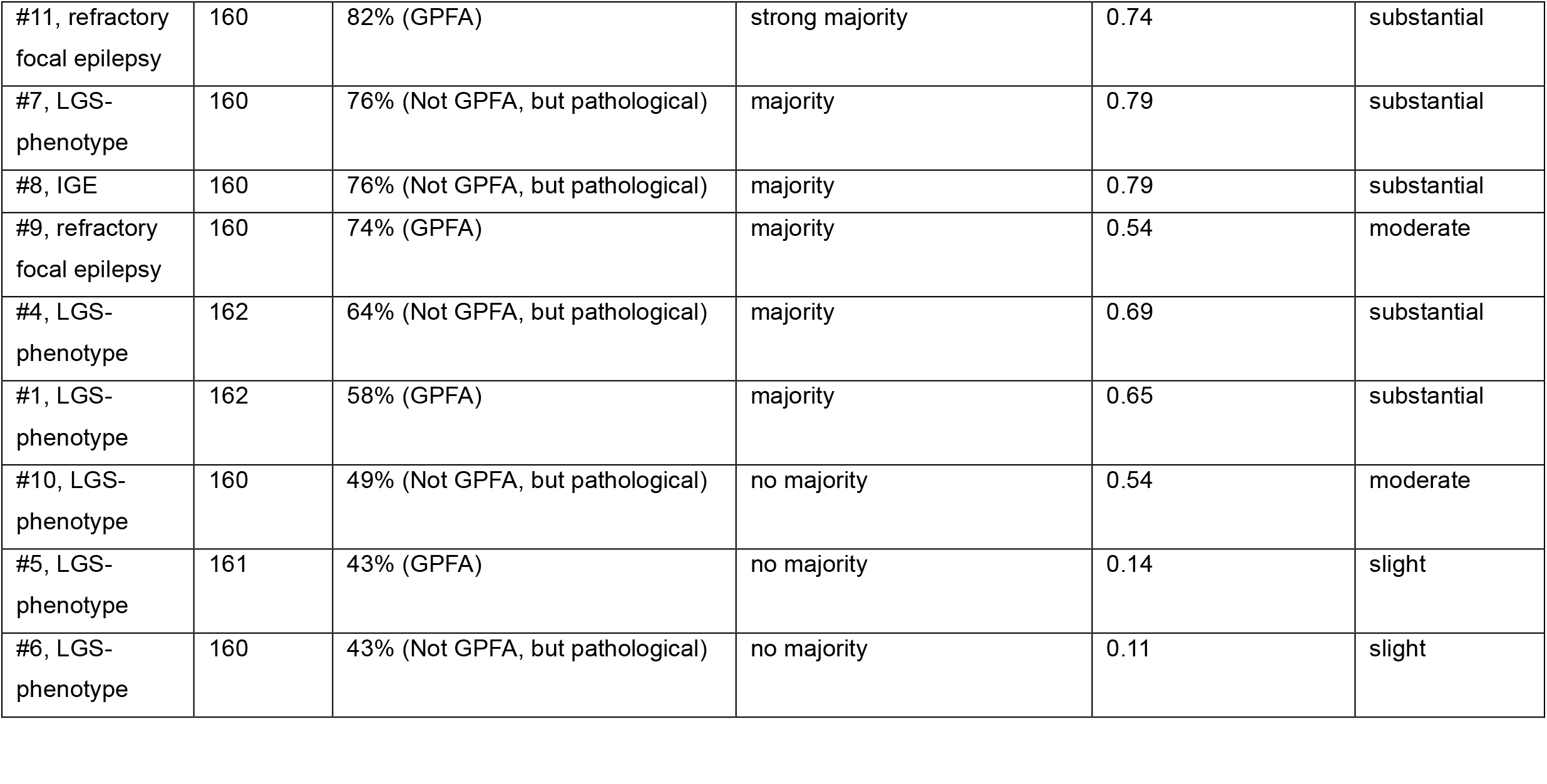
An overview of the agreement found between the EEG clips. Clips are ranked here according to % agreement on the most selected response option. Secondary analysis with Gwet’s AC2 for inter-rater agreement demonstrated broad concordance in ranking, supporting the robustness of the findings.

| EEG clip # | Raters | Primary analysis |  | Secondary analysis |  |
| --- | --- | --- | --- | --- | --- |
|  |  | % agreement (response option) | Majority-ranking | Gwet’s AC2 | Interpretation of agreement |
| #2, non-refractory focal epilepsy | 160 | 88% (not pathological) | strong majority | 0.76 | substantial |
| #3, LGS-phenotype | 162 | 85% (GPFA) | strong majority | 0.70 | substantial |
| #11, refractory focal epilepsy | 160 | 82% (GPFA) | strong majority | 0.74 | substantial |
| #7, LGS-phenotype | 160 | 76% (Not GPFA, but pathological) | majority | 0.79 | substantial |
| #8, IGE | 160 | 76% (Not GPFA, but pathological) | majority | 0.79 | substantial |
| #9, refractory focal epilepsy | 160 | 74% (GPFA) | majority | 0.54 | moderate |
| #4, LGS-phenotype | 162 | 64% (Not GPFA, but pathological) | majority | 0.69 | substantial |
| #1, LGS-phenotype | 162 | 58% (GPFA) | majority | 0.65 | substantial |
| #10, LGS-phenotype | 160 | 49% (Not GPFA, but pathological) | no majority | 0.54 | moderate |
| #5, LGS-phenotype | 161 | 43% (GPFA) | no majority | 0.14 | slight |
| #6, LGS-phenotype | 160 | 43% (Not GPFA, but pathological) | no majority | 0.11 | slight |

### Features of GPFA (*S1*)

87% of respondents considered GPFA to occur in epilepsies other than LGS. Common responses included: focal epilepsy, IGE (specifically “bad” IGE), generalized epilepsies or epileptic encephalopathies, tonic seizures, focal cortical dysplasia, and refractory epilepsies (*S2*).

A strong majority (81%) considered GPFA to have no lower limit for amplitude, provided it stands out from the background activity. 54% considered GPFA to have no criterion for upper limit of frequency. A small majority (57%) agreed that there is no criterion for upper limit of duration, and a similar proportion (54%) considered activity with either anterior or posterior predominance to be acceptable; likewise, 57% deemed lateral predominance acceptable as long as the pattern is present in both hemispheres. There was no majority agreement for lower limit of duration, with 36% selecting a 1.0 second lower limit, and 36% selecting no lower limit. Similarly, the lower limit of frequency lacked majority agreement, with the most common - lower limit of 12 Hz - selected by 30%.

### Primary analysis – percentage agreement for interpretations of 10-second EEG clips

Response distributions to the EEG clips are summarized in *Figure 1* and *S3*. To simplify interpretation, the response options were reclassified into three categories based on percentage agreement:

**Figure 1.**
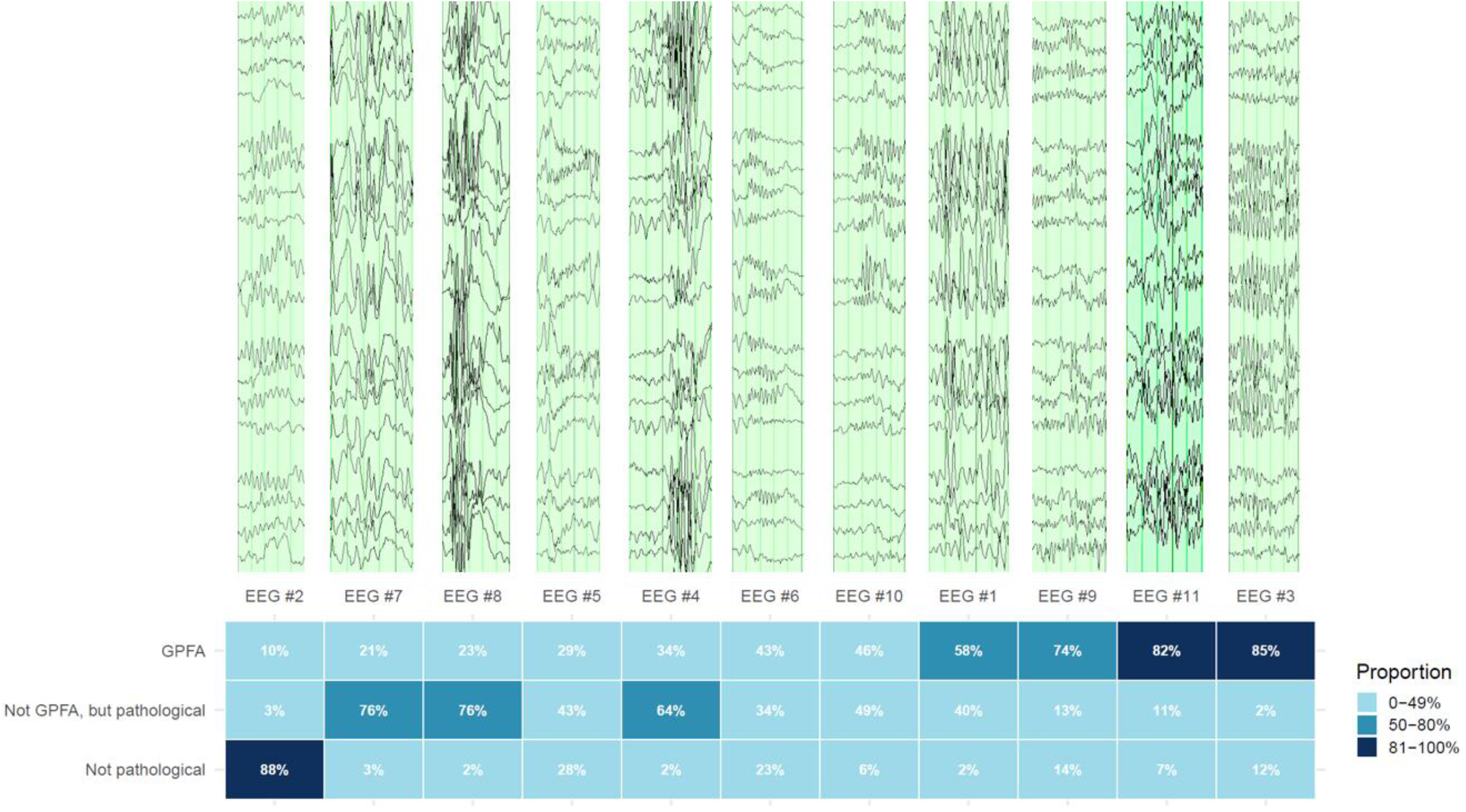
Overview of respondents’ classification of EEG clips, in increasing order (from left to right) of clips most often classified as GPFA. *Note: color-coding is here for illustrative purposes only, and formal assessments of agreement are described in the main text.

1. Strong majority agreement (>80%); #2, #3, #11
2. Small majority agreement (≥50%); #1, #4, #7, #8, #9
3. No majority (<50%); #5, #6, #10

#### EEG clips with strong majority agreement (>80%): #2, #3, #11

EEG clip #2 contained sleep spindles from a patient with non-refractory focal epilepsy; 88% marked this EEG clip as “not pathological”. EEG #3 was from a patient with an LGS-phenotype and EEG #11 was from a patient with refractory focal epilepsy (without tonic seizures); 85% and 82% agreed that this activity was GPFA, respectively.

#### EEG clips with small majority agreement (≥50%): #1, #4, #7, #8, #9

EEGs #1, 4, and 7 were from patients with an LGS-phenotype. For EEG #1, while 58% interpreted this as GPFA, 40% selected “Not GPFA, but pathological” (of which 60% deemed the activity *too high in amplitude* to qualify as GPFA). The type of pathological activity was deemed generalized polyspike trains (GPT) by 46%. For EEG #4, a majority of 64% classified this as “Not GPFA, but pathological” (of which 77% of respondents thought the EEG activity *too short in duration* for GPFA), and the type of pathological activity was considered polyspikes by 64%. Similarly, a majority of 76% interpreted EEG #7 as “Not GPFA, but pathological” (of which 70% deemed the activity *too slow in frequency* for GPFA), with 38% and 36% instead classifying it as either GPT or non-specific burst of diffuse epileptiform activity, respectively.

EEG #8 came from a patient with non-refractory IGE. There was majority agreement on this pattern, with 76% considering it “Not GPFA, but pathological”; 71% considered the activity *too short in duration* for GPFA, and 73% instead interpreted it as polyspikes. EEG #9 was from a patient with refractory focal epilepsy without tonic seizures; 74% considered this GPFA.

#### EEG clips with no majority agreement (<50%): #5, #6, #10

EEG #5 was from a patient with an LGS-phenotype and 43% considered the activity as “Not GPFA, but pathological”, while 28% of respondents selected “Not pathological”. EEG #6 was also from a patient with an LGS-phenotype and was deemed too short to qualify as GPFA by 64% of the respondents who considered it “Not GPFA, but pathological” (34% of total). For EEG #10, also from a patient with an LGS-phenotype, among respondents who judged the activity as “Not GPFA, but pathological” (49%), 77% selected “Other” in the follow-up question, with most citing excessive asymmetry as the reason.

### Secondary analysis – inter-rater agreement on 10-second EEG clips using Gwet’s AC2

Rankings according to percentage agreement and Gwet’s AC2 were largely concordant, with the same five clips ranking highest on both measures (#2, #3, #7, #8, #11), and the same two clips ranking lowest on both measures (#5, #6).

### Subgroup-analyses

Fisher’s exact tests with Holm’s correction for multiple comparisons demonstrated no statistically significant differences in the four groupings studied by experience, pediatric training, geography and caseload (*S4-7*).

## Discussion

This survey sought to evaluate how EEG experts define GPFA, and to assess percentage and inter-rater agreement in the interpretation of fast activity potentially compatible with GPFA. A main finding is that agreement was particularly low for a subset of defining features (lower limits of duration and frequency of GPFA), and highly variable across EEG clips.

87% of respondents considered GPFA to occur in epilepsies other than LGS, with common responses including IGE (with some specifying refractory IGE), patients with tonic seizures, lesional epilepsies and otherwise unspecified epileptic encephalopathies and generalized or refractory epilepsies.

In the literature, GPFA is predominantly, but not exclusively, described in the context of LGS. GPFA and GPT, a morphologically similar EEG pattern, have also been described in IGE, with a possible association with drug resistance and cognitive decline ^23,30–33^. The distinction between GPFA and GPT, however, remains unclear. Sun et al., proposed to distinguish GPT from GPFA based on duration alone, defining GPT as “5 or more rhythmic discharges” lasting from 0.5 to 1.0 seconds, and GPFA as longer than 1.0 seconds in duration ^28^. Alternative minimum durations for GPFA, for either LGS or other epilepsies, include 0.2, 0.25, 0.5, 1.0 and 2.0 seconds ^16,24,33,35,38^. Conrad et al. previously found GPFA and GPT to often co-occur in patients with IGE, with similar morphology on EEG ^27^. Both features correlated with intractable IGE, leading the authors to question the value of an arbitrary duration-informed cut-off; instead, they speculated that GPFA and GPT may be located on an electrographic continuum ^27,28^.

### Duration and frequency of GPFA

About a third of respondents considered a minimum duration of 1.0 seconds for GPFA. Another third selected *no criterion for lower limit*, suggesting that length may not be a primary determinant for differentiating GPFA from similar patterns. However, when asked to interpret an EEG (e.g., clip #8, IGE), most respondents considered the 0.5-second activity too short for GPFA and to instead resemble polyspikes, suggesting that duration remains relevant to a consensus-based definition of GPFA in clinical settings.

57% of respondents selected no upper limit for duration for GPFA; likewise, no consensus exists in the literature, with reported ranges between 1-4 ^14^ and 2-50 seconds ^33^. Similarly, no consensus for tonic seizure duration exists, and a 2023 study of > 10,000 video-EEGs found generalized tonic seizures to last between 2.5 and 50 seconds (median 8.5 seconds)^45^. This potential overlap in duration between GPFA and tonic seizures represents a source of ambiguity, particularly in consideration of their morphological resemblance. In clinical practice, it can be challenging to distinguish GPFAs of longer duration from brief and clinically subtle tonic seizures or electrographic tonic seizures ^35^.

Frequency for GPFA is typically defined as 8-30 Hz, though other ranges have been utilized or reported ^14,16,24,31,33,35^. Consistent with this, we found little agreement on a lower frequency limit, with the most selected answer (12 Hz) only chosen by a third of respondents.

### Amplitude and distribution

A strong majority (81%) selected *no lower limit for amplitude, as long as greater than background*. For regional scalp distribution, a frontal predominance is often described, but a posterior predominance has also been reported ^14^. In this survey, the same opinion was captured: about half of participants considered either anterior or posterior predominance to be acceptable, and another third preferred exclusive anterior predominance. While most respondents considered lateral predominance to be acceptable (provided the pattern is bihemispheric), just under a third allowed a *unilateral pattern in certain clinical contexts*. This suggests that in some cases interpretation of fast paroxysmal activity is dependent on supporting clinical information ^46^.

Only 8% of respondents did not allow regional predominance and only 11% demanded GPFA to be strictly generalized (i.e., involve all channels equally). This aligns with other authors qualifying asymmetric generalizing fast activity in medically refractory localization-related epilepsy as GPFA ^15,47^. A recent study proposed that asymmetric GPFA and tonic seizures may point to a focal driver of widespread epileptic activity in patients progressing from intractable focal epilepsy into an LGS phenotype ^47^.

Together, these findings raise two questions: 1) To what degree is GPFA reporting in daily practice influenced by the clinical information available^46^? and 2) Should GPFA be considered an electrographic entity independent of clinical context, or rather, defined with respect to electroclinical syndromes (e.g. GPFA-LGS, GPFA-IGE) or with clinical context not specified (e.g. GPFA-NOS)?

#### Interpretations of EEG clips

Of the seven clips from patients with LGS phenotypes (EEGs #1, 3, 4, 5, 6, 7, 10), only EEGs #1 and #3 were interpreted as GPFA by a majority of respondents. This limited agreement of GPFA in LGS accords with prior work ^13,46^.

EEG clips #4 and #6 were often deemed “not GPFA, but pathological” due to a short duration of fast activity. In clip #4, 64% agreed on polyspikes, but in EEG clip #6 the final classification of non-GPFA activity was less consistent, suggesting duration is not the leading determinant in separating polyspikes from GPT.

EEG clips #5 and #6, both from patients with LGS-phenotypes, had the lowest agreement. The activity in clip #5 was relatively low-voltage, and clip #6 contained brief ~0.2-0.6-second bursts of fast activity. The LGS-phenotype underlying both these two clips casts doubt on whether an amplitude greater than background and a minimum duration are appropriate requirements for GPFA. In other words, if these patterns constitute only a variant of GPFA, and these patients otherwise meet the clinical criteria for LGS, then perhaps a broader definition of GPFA may be warranted. The substantial phenotypical overlap between clinically-defined LGS (where GPFA documentation may be missing) and electroclinically-defined LGS^38^, suggests that the presence of GPFA may be less central to LGS diagnosis than currently reflected in formal criteria.

Agreement differed substantially for EEG clips #3 and #6, despite being from the same patient, suggesting that multiple fast activity signatures may coexist within the same patient. We acknowledge that 10-second screenshots cannot capture the full spectrum of fast activity in a single patient present in a complete EEG study.

Our EEG samples included two cases of refractory focal epilepsy (clips #9 and #11), and one case of non-refractory IGE, who received majority agreement for GPFA. This suggests that, in the absence of clinical context, the fast activity observed in LGS-phenotypes is not readily distinguishable from that seen in other epilepsies.

#### Strengths and limitations

Our study, due to the granularity of questions, ascertained a limited consensus about GPFA features and permitted identification of specific areas of heterogeneity. The inclusion of a “negative control”, and the fact that the majority correctly identified this activity as spindles, provides reassurance that the survey population were experienced and engaged EEG readers.

This study has limitations. Firstly, due to constraints in outreach efforts, a disproportionate number of respondents originated from the United States and Europe. Hence, responses may reflect geography-specific practices. Selection bias is also inherent to survey-based studies. As previously posited^34^, it is likely that predominantly respondents who consider themselves competent in recognizing LGS or GPFA completed the survey; while this strengthens the credibility of the findings, it also limits generalizability to less specialized clinical neurophysiologists or neurologists. EEG clips were presented as static screenshots and did not permit interaction or adjustment of display settings, which does not reflect clinical practice.

Finally, clinicians were asked to classify single, isolated discharges, whereas in clinical practice multiple discharges would be reviewed from an EEG lasting minutes to hours, and determination on the presence of GPFA would be based on the overall weight of evidence. One consequence of this is that assessment of longer EEG studies may increase agreement among raters.

#### Summary and future directions

There appears to be agreement that a wide variety of paroxysmal fast activity can qualify as GPFA, that GPFA can occur outside of LGS, and that not all fast activity in patients with LGS-phenotypes automatically qualifies as GPFA.

Questions arise from this survey, warranting further study. Is the pathophysiological basis of GPFA in LGS related to, or distinct from, GPT/GPFA as seen in IGE (where tonic seizures do not typically occur) or other epilepsies^27,28^? Is GPFA only an interictal EEG feature, or does it exist on a continuum with tonic seizures ^14,35,48,49^?

We advocate for a common language for GPFA, as inconsistent definitions hamper research into its role as a potential diagnostic or treatment biomarker for LGS, tonic seizures or drug-resistant epilepsy ^11,35,36^. As a next step, we propose a Delphi consensus study for GPFA. Similar to the 2015 ILAE report for classifying status epilepticus ^50^, in the absence of a sufficient understanding of the underlying pathophysiology of GPFA, we argue for an operational definition while ongoing research continues working towards evidence-based criteria (i.e.., biologically-informed or computational, data-driven definitions) ^11,13,38^.

## Supporting information

Original Survey

## Data availability statement

Survey data and code for statistical analysis are available upon reasonable request.

