## Supplemental Tables 1-7 for "Epileptic discharges are classified as generalized paroxysmal fast activity with low expert agreement: *findings from an international survey*"

**Supplemental Material**

**S1.** Defining features of LGS according to survey respondents.

| | Count | Percentage |
|---|---|---|
| **Does GPFA occur in other epilepsies, not limited to LGS?** | | |
| Yes. ______ | 141 | 87% |
| No, it is exclusively found in LGS. | 11 | 7% |
| I am unsure/I do not know. | 10 | 6% |
| **What is the lower limit for duration of GPFA (i.e., the minimum duration for activity to be classified as GPFA)?** | | |
| 0.25 seconds | 6 | 4% |
| 0.5 seconds | 33 | 20% |
| 1.0 seconds | 59 | 36% |
| There is no criterion for lower limit. | 58 | 36% |
| Other ______ | 6 | 4% |
| **What is the upper limit for duration of GPFA (i.e., the maximum duration for activity to be classified as GPFA)?** | | |
| 2 seconds | 2 | 1% |
| 5 seconds | 12 | 7% |
| 10 seconds | 50 | 31% |
| There is no criterion for upper limit. | 92 | 57% |
| Other ______ | 6 | 4% |
| **What is the lower limit for amplitude of GPFA? (i.e., the minimum amplitude for activity to be classified as GPFA?)** | | |
| 50 μV (+/- 25 μV) | 25 | 15% |
| 100 μV (+/- 25 μV) | 3 | 2% |
| There is no lower limit of the amplitude, as long as it is greater relative to background. | 131 | 81% |
| Other ______ | 3 | 2% |
| **What is the lower limit for frequency of GPFA (i.e., the minimum frequency for activity to be classified as GPFA)?** | | |
| 8 Hz | 31 | 19% |
| 10 Hz | 42 | 26% |
| 12 Hz | 48 | 30% |
| There is no criterion for lower limit. | 35 | 22% |
| Other ______ | 6 | 4% |
| **What is the upper limit for frequency of GPFA (i.e., the maximum frequency for activity to be classified as GPFA)?** | | |
| 20 Hz (+/- 5 Hz) | 21 | 13% |
| 30 Hz (+/- 5 Hz) | 49 | 30% |
| There is no criterion for upper limit. | 87 | 54% |
| Other ______ | 5 | 3% |
| **What is the anterior-posterior predominance of GPFA?** | | |
| Anterior predominance | 59 | 36% |

| | | |
|---|---|---|
| Posterior predominance | 2 | 1% |
| Activity with either anterior or posterior predominance is acceptable. | 87 | 54% |
| Regional predominance is not acceptable in GPFA as it is a generalized pattern. | 13 | 8% |
| Other ______ | 1 | 1% |
| **Can GPFA be predominant in one hemisphere or be lateralized to one hemisphere?** | | |
| Yes, an exclusively unilateral pattern can still qualify as GPFA in certain clinical contexts. | 49 | 30% |
| Yes, lateral predominance is acceptable but the pattern has to be present in both hemispheres. | 92 | 57% |
| No, it must be generalized and involve all channels equally. | 18 | 11% |
| Other ______ | 3 | 2% |

**S2.** An overview of respondents' verbatim text-based responses after answering "Yes" to the question: "Does GPFA occur in other epilepsies, not limited to LGS"? Common responses include: IGE (specifically bad "IGE"), generalized epilepsies, epileptic encephalopathies, focal cortical dysplasia, refractory epilepsy and patients with tonic seizures.

| |
|---|
| in rare "idiopathic" generalised epilepsies |
| A few patients with other features of LGS but not all of them |
| all generalized epilepsies |
| Any epileptic encephalopathy |
| Any epileptic encephalopathy |
| Anyone with tonic seizures |
| Atypical idiopathic generalized epilepsy, Doose syndrome |
| autoimmune encephalitis |
| can be see in brain injury, generalized epilepsy |
| Can be see in patients including neonates with brain malformations (lissencephaly as an example) |
| can be seen (or hard to distinguish polyspikes from GPFA) in 'bad IGE' |
| can be seen in patients with tonic seizures |
| Can occur in focal epilepsies as well |
| Can occur in other genetic epilepsies or MCDs while not fulfilling other criteria for LGS |
| Can occur in patients without cognitive impairment |
| cortical dysplasia, rare in EMAS |
| cortical dysplasia, other DEE without SSW |
| DEEs, EE SWAS, some Dravet, some MAE, some focal epilespies (SBS!), and even GGEs! |
| diffuse brain dysfunction |
| dravet, focal epilepsy, progressive myoclonic epilepsy |
| e.g. IESS (as ictal finding) |
| encephalitis, FCD, PMG |
| encephalopathies, generalized epielpsy |
| Epilepsies with tonic seizures |
| epileptic encephalopathies |
| FCD |
| FLE and other focal epilepsies |
| Focal Cortical Dysplasias |
| Focal epilepsies relate to FCD; DEE-SWASS; other genetic DEEs that do not meet criteria for LGS |
| Focal epilepsies with late encephalopathic signs |
| focal epilepsy with mesial frontal onset or occasionally GGE such as JME with atypically long polyspikes |
| For example iess |
| For example, occasionally in EMAtS, and can occasioanlly be seen in other generalized epilepsy syndromes than LGS |
| frontal epilepsy |
| GENERALIZED EPILEPSIES |
| generalized epilepsies |
| Generalized epilepsies with atypical features |

| |
|---|
| generalized epilepsies, encephalitis. etc |
| Generalized epilepsies, Infantile spasms |
| Generalized epilepsy |
| Generalized epilepsy |
| Generalized Epilepsy (particularly drug-resistant, sometimes call intermediate epilepsy9 |
| Generalized epilepsy syndromes other than LGS |
| generalized genetic epilepsy, intermediate syndrome |
| Generalized idiopathic epilepsies that are refractory such as JME |
| genetic epilepsies |
| genetic generalised epilepsy |
| Genetic Generalized Epilepsies, especially refractory |
| genetic generalized epilepsy & lissencephaly |
| GGE |
| GGE |
| GGE, some focal epilepsies ( e.g. parietal) |
| GGE, EE other than LGS |
| GGEs |
| GM-Epilepsy |
| GPFA is a marker of refractoriness in other generalised or focale epilepsies, with high prevalence of mental retardation and multiple seizure types |
| GPFA is a pattern you can find in many epileptic conditions |
| Have rarely seen elsewhere, eg in IGEs (JME) |
| I have seen it in generalized epilepsies |
| idiopathic epilepsy with fast rythms in sleep; some focal structural epilepsies |
| idiopathic generalised epilepsies (a range of these, incl. epilepsies with eyelid myoclonia) |
| IESS or other DEE |
| IESS, any condition where severely disturbed brain by epileptic activity |
| IGE and GGE |
| IGE, patients with tonic seizures of other aetiologies |
| in genetic generalized epilepsy |
| In Genetic Generalized Epilepsy, Other epileptic encepahlopathies |
| In non-specific generalized epilepsies |
| in some refractory IGE and in other GGE with less good outcome, namely eyelid myoclonia with absences, epilepsy with myoclonic absences and perioral myoclonia with absences |
| infantile epileptic spasms syndrome |
| it can occur in Generalised genetic epilepsies |
| it can occur in other epileptic encephalopathies |
| it can occur in other epileptic encephalopathies and/or severe focal epilepsies with focal tonic seizures |
| it is not found only in LGS e.g. other epilepsies with tonic seizures |
| JME (rare but seen) |
| LGS is a electroclinical diagnosis--some people have GPFA without other features of LGS |
| lgs requires additional symptoms/signs |
| Many epilepsies can cause GPFA. Not common though. E.g. dysplasia |

| |
|---|
| More typical of LGS but can be seen in other epilepsies |
| Neonates with epilepsy can have this pattern, as can infants with infantile spasms |
| Non specific DEEs associated with specific genes (i.e. CDKL5-DEE; KCNQ2-DEE) |
| Not all EEGs with GPFA will also have slow spike and wave |
| Occasionally found with IESS, rarely in other situations |
| other DEEs with tonic seizures |
| OTHER DEVELOPMENTAL AND EPILEPTIC ENCEPHALOPATHIES |
| other DRE , focal or generalized |
| other epileptic encephalopathies |
| other epileptic encephalopathies |
| other epileptic encephalopathies not precisely meeting criteria for LGS |
| Other epileptic encephalopathy |
| Other gen DEE |
| other generalised epilepsy syndromes |
| other generalized epilepsies |
| Other generalized epilepsies |
| Other generalized epilepsies, such as EMA can have GPFA |
| Other generalized epilepsy syndromes |
| other generalized epilepsy syndromes |
| Other generaloized epilepsies |
| other nonspecific symptomatic generalized |
| Other structural/metabolic epilepsies |
| other types of generalized epilepsies |
| Patients with DEE |
| Patterns fullfilling criteria for GPFA may appear in focal epilepsies of extratemporal origin, e.g. frontal |
| Poorly controlled primary generalized epilepsy |
| Primary generalized epilepsies |
| progressive myoclonic epilepsies, |
| Rarely but yes another severe generalized epilepsy is not being criteria for LGS |
| rarely can be seen in severe JME/ other frontally predominant epilepsy syndromes- but is rare |
| Seen in other EEGs I read |
| seen in other intractable epilepsies, often generalized, |
| some forms of IGE |
| Some genetic generalised epilepsies or severe drug-resistent epilepsies |
| some GGE |
| Some structural lesions and rarely jeneralize sekondary epilepsies |
| structural abnormality (i have definitely seen it in non-LGS patients) |
| symptomatic generalised epilepsies |
| thinking about the wide range of genetic DEEs that may or may not present as LGS |
| Tonic seizures |
| Tuberous sclerosis and Focal Cortical dysplasias |
| Unspecified epilepsies which do not meet LGS criteria |

| |
|---|
| Various genetic and metabolic disorders. |
| very rarely |
| Yes, in some DEEs, but if there are no tonic seizures, the appropriate age of onset, and the slow spike-wave pattern in wakefulness, it does not qualify as Lennox-Gastaut. |

**S3.** Summary of survey respondents' interpretations of EEG clips. Note, per EEG excerpt, percentages for parent categories are calculated based on number of non-missing responses. E.g., for EEG excerpt #2, 16/160 (rather than 16/162) was used to calculate the percentage of respondents that selected "Yes, this is GPFA".

| | **Count** | **Percentage** |
|---|---|---|
| **EEG excerpt #1** | | |
| **1. Yes, this is GPFA** | 94 | 58% |
| **2. No, this is not GPFA but it is pathological (please see two follow-up questions below)** | 65 | 40% |
| **2a. It does not meet criteria for GPFA, because (check all that apply):** | | |
| Too long duration | 1 | 2% |
| Too short duration | 14 | 22% |
| Too fast frequency | 4 | 6% |
| Too slow frequency | 18 | 28% |
| Too high amplitude | 39 | 60% |
| Too low amplitude | 1 | 2% |
| Other ______ | 5 | 8% |
| **2b. I would classify this pattern instead as (pick one):** | | |
| Polyspikes | 21 | 32% |
| Generalized polyspike train (GPT) | 30 | 46% |
| Non-specific burst of diffuse epileptiform activity | 12 | 19% |
| Other ______ | 2 | 3% |
| **3. No, this is not GPFA and it is not a pathological pattern (e.g., spindles, diffuse beta-activity, arousal)** | 3 | 2% |
| **4. No response** | 0 | 0% |
| **EEG excerpt #2** | | |
| **1. Yes, this is GPFA** | 16 | 10% |
| **2. No, this is not GPFA but it is pathological (please see two follow-up questions below)** | 4 | 3% |
| **2a. It does not meet criteria for GPFA, because (check all that apply):** | | |
| Too long duration | 0 | 0% |
| Too short duration | 1 | 25% |
| Too fast frequency | 0 | 0% |

| | | |
|---|---|---|
| Too slow frequency | 1 | 25% |
| Too high amplitude | 0 | 0% |
| Too low amplitude | 2 | 50% |
| Other ______ | 3 | 75% |
| **2b. I would classify this pattern instead as (pick one):** | | |
| Polyspikes | 1 | 25% |
| Generalized polyspike train (GPT) | 0 | 0% |
| Non-specific burst of diffuse epileptiform activity | 0 | 0% |
| Other ______ | 3 | 75% |
| **3. No, this is not GPFA and it is not a pathological pattern (e.g., spindles, diffuse beta-activity, arousal)** | 140 | 88% |
| **4. No response** | 2 | 1% |
| **EEG excerpt #3** | | |
| **1. Yes, this is GPFA** | 138 | 85% |
| **2. No, this is not GPFA but it is pathological (please see two follow-up questions below)** | 4 | 2% |
| **2a. It does not meet criteria for GPFA, because (check all that apply):** | | |
| Too long duration | 1 | 25% |
| Too short duration | 0 | 0% |
| Too fast frequency | 1 | 25% |
| Too slow frequency | 0 | 0% |
| Too high amplitude | 1 | 25% |
| Too low amplitude | 0 | 0% |
| Other ______ | 2 | 50% |
| **2b. I would classify this pattern instead as (pick one):** | | |
| Polyspikes | 0 | 0% |
| Generalized polyspike train (GPT) | 0 | 0% |
| Non-specific burst of diffuse epileptiform activity | 4 | 100% |
| Other ______ | 0 | 0% |
| **3. No, this is not GPFA and it is not a pathological pattern (e.g., spindles, diffuse beta-activity, arousal)** | 20 | 12% |
| **4. No response** | 0 | 0% |
| **EEG excerpt #4** | | |
| **1. Yes, this is GPFA** | 55 | 34% |
| **2. No, this is not GPFA but it is pathological (please see two follow-up questions below)** | 103 | 64% |
| **2a. It does not meet criteria for GPFA, because (check all that apply):** | | |
| Too long duration | 0 | 0% |

| | | |
|---|---|---|
| Too short duration | 79 | 77% |
| Too fast frequency | 3 | 3% |
| Too slow frequency | 2 | 2% |
| Too high amplitude | 22 | 21% |
| Too low amplitude | 0 | 0% |
| Other ______ | 15 | 15% |
| **2b. I would classify this pattern instead as (pick one):** | | |
| Polyspikes | 66 | 64% |
| Generalized polyspike train (GPT) | 23 | 22% |
| Non-specific burst of diffuse epileptiform activity | 10 | 10% |
| Other ______ | 4 | 4% |
| **3. No, this is not GPFA and it is not a pathological pattern (e.g., spindles, diffuse beta-activity, arousal)** | 4 | 2% |
| **4. No response** | 0 | 0% |
| **EEG excerpt #5** | | |
| **1. Yes, this is GPFA** | 47 | 29% |
| **2. No, this is not GPFA but it is pathological (please see two follow-up questions below)** | 69 | 43% |
| **2a. It does not meet criteria for GPFA, because (check all that apply):** | | |
| Too long duration | 2 | 3% |
| Too short duration | 24 | 35% |
| Too fast frequency | 14 | 20% |
| Too slow frequency | 2 | 3% |
| Too high amplitude | 0 | 0% |
| Too low amplitude | 16 | 23% |
| Other ______ | 35 | 51% |
| **2b. I would classify this pattern instead as (pick one):** | | |
| Polyspikes | 15 | 22% |
| Generalized polyspike train (GPT) | 4 | 6% |
| Non-specific burst of diffuse epileptiform activity | 29 | 42% |
| Other ______ | 21 | 30% |
| **3. No, this is not GPFA and it is not a pathological pattern (e.g., spindles, diffuse beta-activity, arousal)** | 45 | 28% |
| **4. No response** | 1 | 1% |
| **EEG excerpt #6** | | |
| **1. Yes, this is GPFA** | 69 | 43% |
| **2. No, this is not GPFA but it is pathological (please see two follow-up questions below)** | 55 | 34% |

| | | |
|---|---|---|
| **2a. It does not meet criteria for GPFA, because (check all that apply):** | | |
| Too long duration | 2 | 4% |
| Too short duration | 35 | 64% |
| Too fast frequency | 2 | 4% |
| Too slow frequency | 1 | 2% |
| Too high amplitude | 0 | 0% |
| Too low amplitude | 5 | 9% |
| Other ______ | 16 | 29% |
| **2b. I would classify this pattern instead as (pick one):** | | |
| Polyspikes | 15 | 27% |
| Generalized polyspike train (GPT) | 15 | 27% |
| Non-specific burst of diffuse epileptiform activity | 14 | 25% |
| Other ______ | 11 | 20% |
| **3. No, this is not GPFA and it is not a pathological pattern (e.g., spindles, diffuse beta-activity, arousal)** | 36 | 23% |
| **4. No response** | 2 | 1% |
| **EEG excerpt #7** | | |
| **1. Yes, this is GPFA** | 34 | 21% |
| **2. No, this is not GPFA but it is pathological (please see two follow-up questions below)** | 121 | 76% |
| **2a. It does not meet criteria for GPFA, because (check all that apply):** | | |
| Too long duration | 7 | 6% |
| Too short duration | 3 | 2% |
| Too fast frequency | 1 | 1% |
| Too slow frequency | 85 | 70% |
| Too high amplitude | 27 | 22% |
| Too low amplitude | 1 | 1% |
| Other ______ | 19 | 16% |
| **2b. I would classify this pattern instead as (pick one):** | | |
| Polyspikes | 25 | 21% |
| Generalized polyspike train (GPT) | 46 | 38% |
| Non-specific burst of diffuse epileptiform activity | 43 | 36% |
| Other ______ | 6 | 5% |
| **3. No, this is not GPFA and it is not a pathological pattern (e.g., spindles, diffuse beta-activity, arousal)** | 5 | 3% |
| **4. No response** | 2 | 1% |
| **EEG excerpt #8** | | |
| **1. Yes, this is GPFA** | 36 | 23% |

| | | |
|---|---|---|
| **2. No, this is not GPFA but it is pathological (please see two follow-up questions below)** | 121 | 76% |
| **2a. It does not meet criteria for GPFA, because (check all that apply):** | | |
| Too long duration | 0 | 0% |
| Too short duration | 86 | 71% |
| Too fast frequency | 6 | 5% |
| Too slow frequency | 1 | 1% |
| Too high amplitude | 32 | 26% |
| Too low amplitude | 0 | 0% |
| Other ______ | 16 | 13% |
| **2b. I would classify this pattern instead as (pick one):** | | |
| Polyspikes | 88 | 73% |
| Generalized polyspike train (GPT) | 20 | 17% |
| Non-specific burst of diffuse epileptiform activity | 8 | 7% |
| Other ______ | 4 | 3% |
| **3. No, this is not GPFA and it is not a pathological pattern (e.g., spindles, diffuse beta-activity, arousal)** | 3 | 2% |
| **4. No response** | 2 | 1% |
| **EEG excerpt #9** | | |
| **1. Yes, this is GPFA** | 118 | 74% |
| **2. No, this is not GPFA but it is pathological (please see two follow-up questions below)** | 20 | 13% |
| **2a. It does not meet criteria for GPFA, because (check all that apply):** | | |
| Too long duration | 4 | 20% |
| Too short duration | 0 | 0% |
| Too fast frequency | 2 | 10% |
| Too slow frequency | 1 | 5% |
| Too high amplitude | 0 | 0% |
| Too low amplitude | 4 | 20% |
| Other ______ | 10 | 50% |
| **2b. I would classify this pattern instead as (pick one):** | | |
| Polyspikes | 4 | 20% |
| Generalized polyspike train (GPT) | 2 | 10% |
| Non-specific burst of diffuse epileptiform activity | 10 | 50% |
| Other ______ | 4 | 20% |
| **3. No, this is not GPFA and it is not a pathological pattern (e.g., spindles, diffuse beta-activity, arousal)** | 22 | 14% |
| **4. No response** | 2 | 1% |

| | | |
|---|---|---|
| **EEG excerpt #10** | | |
| **1. Yes, this is GPFA** | 73 | 46% |
| **2. No, this is not GPFA but it is pathological (please see two follow-up questions below)** | 78 | 49% |
| **2a. It does not meet criteria for GPFA, because (check all that apply):** | | |
| Too long duration | 1 | 1% |
| Too short duration | 12 | 15% |
| Too fast frequency | 3 | 4% |
| Too slow frequency | 2 | 3% |
| Too high amplitude | 2 | 3% |
| Too low amplitude | 0 | 0% |
| Other ______ | 60 | 77% |
| **2b. I would classify this pattern instead as (pick one):** | | |
| Polyspikes | 36 | 46% |
| Generalized polyspike train (GPT) | 8 | 10% |
| Non-specific burst of diffuse epileptiform activity | 16 | 21% |
| Other ______ | 18 | 23% |
| **3. No, this is not GPFA and it is not a pathological pattern (e.g., spindles, diffuse beta-activity, arousal)** | 9 | 6% |
| **4. No response** | 2 | 1% |
| **EEG excerpt #11** | | |
| **1. Yes, this is GPFA** | 131 | 82% |
| **2. No, this is not GPFA but it is pathological (please see two follow-up questions below)** | 18 | 11% |
| **2a. It does not meet criteria for GPFA, because (check all that apply):** | | |
| Too long duration | 1 | 6% |
| Too short duration | 0 | 0% |
| Too fast frequency | 5 | 28% |
| Too slow frequency | 0 | 0,0% |
| Too high amplitude | 2 | 11% |
| Too low amplitude | 0 | 0% |
| Other ______ | 12 | 67% |
| **2b. I would classify this pattern instead as (pick one):** | | |
| Polyspikes | 2 | 11% |
| Generalized polyspike train (GPT) | 5 | 28% |
| Non-specific burst of diffuse epileptiform activity | 10 | 56% |
| Other ______ | 1 | 6% |
| **3. No, this is not GPFA and it is not a pathological pattern (e.g., spindles, diffuse beta-activity, arousal)** | 11 | 7% |
| **4. No response** | 2 | 1% |

**Supplemental Tables 4-7 (S4-7)**

**S4.** Fisher-exact tests with Holm's correction for multiple comparisons (**adjusted p-value**) to explore subgroup differences in the distribution of responses to the 10-second EEG clips for clinicians with 10 or fewer years vs. more than 10 years of clinical experience. Statistical significance was evaluated at α = 0.05.

| EEG clip | p-value | adjusted p-value | significant |
|---|---|---|---|
| #1 | 0.191 | 1 | false |
| #2 | 0.782 | 1 | false |
| #3 | 0.552 | 1 | false |
| #4 | 0.949 | 1 | false |
| #5 | 0.069 | 0.755 | false |
| #6 | 0.392 | 1 | false |
| #7 | 0.802 | 1 | false |
| #8 | 0.195 | 1 | false |
| #9 | 0.772 | 1 | false |
| #10 | 0.223 | 1 | false |
| #11 | 0.164 | 1 | false |

**S5.** Fisher-exact tests with Holm's correction for multiple comparisons (**adjusted p-value**) to explore subgroup differences in the distribution of responses to the 10-second EEG clips for child vs. adult neurologists. Statistical significance was evaluated at α = 0.05.

| EEG clip | p-value | adjusted p-value | significant |
|---|---|---|---|
| #1 | 0.124 | 1 | false |
| #2 | 0.719 | 1 | false |
| #3 | 0.258 | 1 | false |
| #4 | 1.00 | 1 | false |
| #5 | 0.125 | 1 | false |
| #6 | 0.463 | 1 | false |
| #7 | 0.842 | 1 | false |
| #8 | 0.148 | 1 | false |
| #9 | 0.926 | 1 | false |
| #10 | 0.329 | 1 | false |
| #11 | 0.154 | 1 | false |

**S6**. Fisher-exact tests with Holm's correction for multiple comparisons (**adjusted p-value**) to explore subgroup differences in the distribution of responses to the 10-second EEG clips for European vs. North American respondents. Statistical significance was evaluated at α = 0.05.

| EEG clip | p-value | adjusted p-value | significant |
|---|---|---|---|
| #1 | 0.132 | 1 | false |
| #2 | 1.000 | 1 | false |
| #3 | 0.671 | 1 | false |
| #4 | 0.269 | 1 | false |
| #5 | 0.821 | 1 | false |
| #6 | 0.191 | 1 | false |
| #7 | 0.348 | 1 | false |
| #8 | 0.438 | 1 | false |
| #9 | 0.241 | 1 | false |
| #10 | 0.092 | 1 | false |
| #11 | 0.528 | 1 | false |

**S7**. Fisher-exact tests with Holm's correction for multiple comparisons (**adjusted p-value**) to explore subgroup differences in the distribution of responses to the 10-second EEG clips for caseload (0-25 vs. 25-50 LGS patients per year). Statistical significance was evaluated at α = 0.05.

| EEG clip | p-value | adjusted p-value | significant |
|---|---|---|---|
| #1 | 0.398 | 1 | false |
| #2 | 0.616 | 1 | false |
| #3 | 0.929 | 1 | false |
| #4 | 0.902 | 1 | false |
| #5 | 0.520 | 1 | false |
| #6 | 0.521 | 1 | false |
| #7 | 0.139 | 1 | false |
| #8 | 0.940 | 1 | false |
| #9 | 0.099 | 1 | false |
| #10 | 0.187 | 1 | false |
| #11 | 0.320 | 1 | false |
