## Supplementary material for "Epileptic discharges are classified as generalized paroxysmal fast activity with low expert agreement: *findings from an international survey*": Original Survey

PLEASE READ For certain images, the quality may appear degraded when viewed on a PC. To resolve this, simply right-clik on the image and select "Open image in new tab". This should display the image in its original quality.

---

The exact features considered necessary for diagnosis of Lennox-Gastaut syndrome (LGS) have evolved over time, and are variably applied by clinicians [1-5].

In the 2022 ILAE guidelines, generalized paroxysmal fast activity (GPFA) is a mandatory component for diagnosis of LGS. It can be present in either current or historical patient EEGs [6]. GPFA's generalized EEG pattern, its presence in patients with tonic seizures irrespective of etiology, and its morphological overlap with the EEG pattern of tonic seizures suggest a central role in epileptic networks in LGS [7,8]. A universal definition, however, is currently lacking.

The literature on what constitutes GPFA is inconsistent. The morphological, electrophysiological, and temporal (e.g., minimum duration) characteristics are defined differently across studies.

The lack of a uniform definition hampers progress in LGS research. This includes research into the role of GPFA as a potential biomarker of tonic seizures and response to therapy [7,9].

Here, we aim to collect your opinion on GPFA and its EEG mimics to identify areas of agreement and disagreement. We kindly ask medical professionals in the field of neurology, clinical neurophysiology, and epilepsy to fill out the following questionnaire.

Thank you for your participation. Please reach out with questions.

Sem Kampman, Utrecht University

Jurriaan Peters, Boston Children's Hospital

PS Please feel free to forward to your colleagues!

---

References[1] Rijckevorsel K van. Treatment of Lennox-Gastaut syndrome: overview and recent findings. Neuropsychiatr Dis Treat 2008;4:1001. https://doi.org/10.2147/NDT.S1668.

[2] Camfield PR. Definition and natural history of Lennox-Gastaut syndrome. Epilepsia 2011;52:3-9. https://doi.org/10.1111/J.1528-1167.2011.03177.X.

[3] Panayiotopoulos C. Epileptic Encephalopathies in Infancy and Early Childhood in Which the Epileptiform Abnormalities May Contribute to Progressive Dysfunction 2005.

1 What is your clinical experience?

◯ Adult neurology resident or epilepsy fellow/trainee
◯ Adult neurologist, staff physician (< 5 years experience)
◯ Adult neurologist, staff physician (5-10 years experience)
◯ Adult neurologist, staff physician (>10 years experience)
◯ Child neurology resident or epilepsy fellow/trainee
◯ Child neurologist, staff physician (< 5 years experience)
◯ Child neurologist, staff physician (5-10 years experience)
◯ Child neurologist, staff physician (>10 years experience)
◯ Other ______

2 Do you consider yourself an EEG expert?

◯ Yes (e.g. dedicated training, board certification in epilepsy or clinical neurophysiology, regular EEG interpretation in clinical setting)
◯ No

3 In which healthcare setting do you currently work?

◯ University-based hospital / academic hospital
◯ Non university-based hospital
◯ Private practice
◯ Other ______

4 In which country do you currently work?

__________________________________

5 How many patients with LGS do you see per year?

◯ < 10
◯ 10 - 25
◯ 25 - 50
◯ > 50

6 Does GPFA occur in other epilepsies, not limited to LGS?

◯ Yes. ______
◯ No, it is exclusively found in LGS.
◯ I am unsure/I do not know.

7 What is the lower limit for duration of GPFA (i.e., the minimum duration for activity to be classified as GPFA)?

◯ 0.25 seconds
◯ 0.5 seconds
◯ 1.0 seconds
◯ There is no criterion for lower limit.
◯ Other ______

8 What is the upper limit for duration of GPFA (i.e., the maximum duration for activity to be classified as GPFA)?

◯ 2 seconds
◯ 5 seconds
◯ 10 seconds
◯ There is no criterion for upper limit.
◯ Other ______

9 What is the lower limit for amplitude of GPFA? (i.e., the minimum amplitude for activity to be classified as GPFA?)

◯ 50 µV (+/- 25 µV)
◯ 100 µV (+/- 25 µV)
◯ There is no lower limit of the amplitude, as long as it is greater relative to background.
◯ Other ______

10 What is the lower limit for frequency of GPFA (i.e., the minimum frequency for activity to be classified as GPFA)?

◯ 8 Hz
◯ 10 Hz
◯ 12 Hz
◯ There is no criterion for lower limit.
◯ Other ______

11 What is the upper limit for frequency of GPFA (i.e., the maximum frequency for activity to be classified as GPFA)?

◯ 20 Hz (+/- 5 Hz)
◯ 30 Hz (+/- 5 Hz)
◯ There is no criterion for upper limit.
◯ Other ______

12 What is the anterior-posterior predominance of GPFA?

◯ Anterior predominance
◯ Posterior predominance
◯ Activity with either anterior or posterior predominance is acceptable.
◯ Regional predominance is not acceptable in GPFA as it is a generalized pattern.
◯ Other ______

13 Can GPFA be predominant in one hemisphere or be lateralized to one hemisphere?

◯ Yes, an exclusively unilateral pattern can still qualify as GPFA in certain clinical contexts.
◯ Yes, lateral predominance is acceptable but the pattern has to be present in both hemispheres.
◯ No, it must be generalized and involve all channels equally.
◯ Other ______

1- Would you classify the following selected EEG fragment as GPFA? (same fragment, shown using two different sensitivities)

◯ Yes, this is GPFA
◯ No, this is not GPFA but it is pathological (please see two follow-up questions below)
◯ No, this is not GPFA and it is not a pathological pattern (e.g., spindles, diffuse beta-activity, arousal)

______ ______
Female, 10 years old

It does not meet criteria for GPFA, because (check all that apply):

☐ Too long duration
☐ Too short duration
☐ Too fast frequency
☐ Too slow frequency
☐ Too high amplitude
☐ Too low amplitude
☐ Other ______

I would classify this pattern instead as (pick one):

◯ Polyspikes
◯ Generalized polyspike train (GPT)
◯ Non-specific burst of diffuse epileptiform activity
◯ Other ______

2- Would you classify the following selected EEG fragment as GPFA? (same fragment, shown using two different sensitivities)

◯ Yes, this is GPFA
◯ No, this is not GPFA but it is pathological (please see two follow-up questions below)
◯ No, this is not GPFA and it is not a pathological pattern (e.g., spindles, diffuse beta-activity, arousal)

______ ______
Female, 8 years old

It does not meet criteria for GPFA, because (check all that apply):

☐ Too long duration
☐ Too short duration
☐ Too fast frequency
☐ Too slow frequency
☐ Too high amplitude
☐ Too low amplitude
☐ Other ______

I would classify this pattern instead as (pick one):

◯ Polyspikes
◯ Generalized polyspike train (GPT)
◯ Non-specific burst of diffuse epileptiform activity
◯ Other ______

3- Would you classify the following selected EEG fragment as GPFA? (same fragment, shown using two different sensitivities)

◯ Yes, this is GPFA
◯ No, this is not GPFA but it is pathological (please see two follow-up questions below)
◯ No, this is not GPFA and it is not a pathological pattern (e.g., spindles, diffuse beta-activity, arousal)

______ ______
Female, 15 years old

It does not meet criteria for GPFA, because (check all that apply):

☐ Too long duration
☐ Too short duration
☐ Too fast frequency
☐ Too slow frequency
☐ Too high amplitude
☐ Too low amplitude
☐ Other ______

I would classify this pattern instead as (pick one):

◯ Polyspikes
◯ Generalized polyspike train (GPT)
◯ Non-specific burst of diffuse epileptiform activity
◯ Other ______

4- Would you classify the following selected EEG fragment as GPFA? (same fragment, shown using two different sensitivities)

◯ Yes, this is GPFA
◯ No, this is not GPFA but it is pathological (please see two follow-up questions below)
◯ No, this is not GPFA and it is not a pathological pattern (e.g., spindles, diffuse beta-activity, arousal)

______ ______
Male, 9 years old

It does not meet criteria for GPFA, because (check all that apply):

☐ Too long duration
☐ Too short duration
☐ Too fast frequency
☐ Too slow frequency
☐ Too high amplitude
☐ Too low amplitude
☐ Other ______

I would classify this pattern instead as (pick one):

◯ Polyspikes
◯ Generalized polyspike train (GPT)
◯ Non-specific burst of diffuse epileptiform activity
◯ Other ______

5- Would you classify the following selected EEG fragment as GPFA? (same fragment, shown using two different sensitivities)

◯ Yes, this is GPFA
◯ No, this is not GPFA but it is pathological (please see two follow-up questions below)
◯ No, this is not GPFA and it is not a pathological pattern (e.g., spindles, diffuse beta-activity, arousal)

______ ______
Female, 11 years old

It does not meet criteria for GPFA, because (check all that apply):

☐ Too long duration
☐ Too short duration
☐ Too fast frequency
☐ Too slow frequency
☐ Too high amplitude
☐ Too low amplitude
☐ Other ______

I would classify this pattern instead as (pick one):

◯ Polyspikes
◯ Generalized polyspike train (GPT)
◯ Non-specific burst of diffuse epileptiform activity
◯ Other ______

6- Would you classify the following selected EEG fragment as GPFA? (same fragment, shown using two different sensitivities)

◯ Yes, this is GPFA
◯ No, this is not GPFA but it is pathological (please see two follow-up questions below)
◯ No, this is not GPFA and it is not a pathological pattern (e.g., spindles, diffuse beta-activity, arousal)

______ ______
Female, 15 years old

It does not meet criteria for GPFA, because (check all that apply):

☐ Too long duration
☐ Too short duration
☐ Too fast frequency
☐ Too slow frequency
☐ Too high amplitude
☐ Too low amplitude
☐ Other ______

I would classify this pattern instead as (pick one):

◯ Polyspikes
◯ Generalized polyspike train (GPT)
◯ Non-specific burst of diffuse epileptiform activity
◯ Other ______

7- Would you classify the following selected EEG fragment as GPFA? (same fragment, shown using two different sensitivities)

◯ Yes, this is GPFA
◯ No, this is not GPFA but it is pathological (please see two follow-up questions below)
◯ No, this is not GPFA and it is not a pathological pattern (e.g., spindles, diffuse beta-activity, arousal)

______ ______
Female, 8 years old

It does not meet criteria for GPFA, because (check all that apply):

☐ Too long duration
☐ Too short duration
☐ Too fast frequency
☐ Too slow frequency
☐ Too high amplitude
☐ Too low amplitude
☐ Other ______

I would classify this pattern instead as (pick one):

◯ Polyspikes
◯ Generalized polyspike train (GPT)
◯ Non-specific burst of diffuse epileptiform activity
◯ Other ______

8- Would you classify the following selected EEG fragment as GPFA? (same fragment, shown using two different sensitivities)

◯ Yes, this is GPFA
◯ No, this is not GPFA but it is pathological (please see two follow-up questions below)
◯ No, this is not GPFA and it is not a pathological pattern (e.g., spindles, diffuse beta-activity, arousal)

______ ______
Female, 16 years old

It does not meet criteria for GPFA, because (check all that apply):

☐ Too long duration
☐ Too short duration
☐ Too fast frequency
☐ Too slow frequency
☐ Too high amplitude
☐ Too low amplitude
☐ Other ______

I would classify this pattern instead as (pick one):

◯ Polyspikes
◯ Generalized polyspike train (GPT)
◯ Non-specific burst of diffuse epileptiform activity
◯ Other ______

9- Would you classify the following selected EEG fragment as GPFA? (same fragment, shown using two different sensitivities)

◯ Yes, this is GPFA
◯ No, this is not GPFA but it is pathological (please see two follow-up questions below)
◯ No, this is not GPFA and it is not a pathological pattern (e.g., spindles, diffuse beta-activity, arousal)

______ ______
Female, 16 years old

It does not meet criteria for GPFA, because (check all that apply):

☐ Too long duration
☐ Too short duration
☐ Too fast frequency
☐ Too slow frequency
☐ Too high amplitude
☐ Too low amplitude
☐ Other ______

I would classify this pattern instead as (pick one):

◯ Polyspikes
◯ Generalized polyspike train (GPT)
◯ Non-specific burst of diffuse epileptiform activity
◯ Other ______

10- Would you classify the following selected EEG fragment as GPFA? (same fragment, shown using two different sensitivities)

◯ Yes, this is GPFA
◯ No, this is not GPFA but it is pathological (please see two follow-up questions below)
◯ No, this is not GPFA and it is not a pathological pattern (e.g., spindles, diffuse beta-activity, arousal)

______ ______
Female, 16 years old

It does not meet criteria for GPFA, because (check all that apply):

☐ Too long duration
☐ Too short duration
☐ Too fast frequency
☐ Too slow frequency
☐ Too high amplitude
☐ Too low amplitude
☐ Other ______

I would classify this pattern instead as (pick one):

◯ Polyspikes
◯ Generalized polyspike train (GPT)
◯ Non-specific burst of diffuse epileptiform activity
◯ Other ______

11- Would you classify the following selected EEG fragment as GPFA? (same fragment, shown using two different sensitivities)

◯ Yes, this is GPFA
◯ No, this is not GPFA but it is pathological (please see two follow-up questions below)
◯ No, this is not GPFA and it is not a pathological pattern (e.g., spindles, diffuse beta-activity, arousal)

______ ______
Male, 26 years old

It does not meet criteria for GPFA, because (check all that apply):

☐ Too long duration
☐ Too short duration
☐ Too fast frequency
☐ Too slow frequency
☐ Too high amplitude
☐ Too low amplitude
☐ Other ______

I would classify this pattern instead as (pick one):

◯ Polyspikes
◯ Generalized polyspike train (GPT)
◯ Non-specific burst of diffuse epileptiform activity
◯ Other ______
